# Socioeconomic Inequalities in Adult Mortality across Small Areas in Brazil: Exploration Analysis Using Machine Learning Models

**DOI:** 10.1101/2020.08.15.20175687

**Authors:** Natália Martins Arruda, Tiago José de Carvalho, Luciana Correia Alves

## Abstract

Future progress in life expectancy in all countries depends, to some degree, on a reduction in adult mortality. Significant regional differences are found in Brazil, with high adult mortality rates in some areas. The aim of the present study was to investigate associations between the probability of adult deaths in microregions of Brazil in 2010 and socioeconomic, structural, contextual and health-related factors. The analyses were based on data from the 2010 demographic census and the Mortality Information System. The machine learning model was used to establish determinants of the probability of adult deaths. Machine learning methods have considerable potential for this type of analysis, enabling a better understanding of the interactions among different factors. The algorithms employed (*Random Forest, Extreme Gradient Boosted Trees* and *Support Vector Machine*) obtained a good performance and proved to be effective at analyzing the variables and correlations with the outcome (probability of adult death). The results showed the mortality due to external causes, the employment rate, sex ratio, vaccine coverage, proportion of blacks, poverty rate, aging index and proportion of whites had the best predictive power regarding the probability of adult deaths using the algorithm with the best performance (*Extreme Gradient Boosted Trees*).

## Introduction

Socioeconomic inequalities play an important role in the understanding of the accelerated decline in mortality in developing countries. For instance, Santos and Noronha (2001) showed that economically privileged groups have lower all-cause mortality. The decline in the mortality rate is greater in populations with more schooling as well as better housing and sanitation conditions (Kunitz 1987). According to Cutler, Deaton and Lleras-Muney (2006), individuals with a low income, low education and low social status often die at a younger age compared to those with more schooling and a higher income.

In Brazil, there has been an overall reduction in the risk of morality in children, but this has been replaced with an increased risk beginning at 15 years of age, which is the onset of the transition to adulthood (Silva et al. 2016). Therefore, the investigation of adult mortality, its geographic distribution and its association with socioeconomic characteristics is essential to understating contextual factors, causes and consequences regarding the generation of health inequalities in Brazil (Rentería-Peréz and Turra 2008; Pereira and Queiroz 2016).

Socioeconomic inequalities in the context child mortality have been widely investigated in Brazil and the world (Mosley and Chen 1984; Victora et al. 2000; Castro and Simões 2009). In contrast, most studies addressing adult mortality have focused on the quality of the data and coverage rather than factors of inequality (Moura et al. 2016; Walque and Wilmer 2013; Vasconcelos and França 2012). The determinants of adult mortality are explored little in Brazil (e.g., Queiroz et al. 2017).

According to Queiroz et al. (2017), there was considerable coverage of deaths between 1980 and 2010 in studies analyzing the quality of data related to adult mortality. The authors found that adult mortality in meso-regions of Brazil converges among the regions of the country in the period analyzed. Moreover, significant differences were found, with some areas of high mortality. Reductions in mortality rates in areas with a low socioeconomic status may indicate better living conditions.

The identification of characteristics on the household and regional levels that exert an impact on the risk of adult mortality can assist in the development of policies and actions directed at this health and socioeconomic problem. Future progress in life expectancy will, to some degree, depend on the reduction in adult mortality rates (Vallin and Meslé, 2004).

Contextual factors are important determinants of adult mortality. After considering individual characteristics, such as sex, age, race and education, the government plays an important role in increasing or reducing the mortality rate in this age group. In terms of schooling, Montez et al. (2019) revealed that public policies exert a greater impact on adults with low schooling and income, arguing that adults with a higher level of schooling can improve their health and longevity with their own resources independently of public policies addressing unemployment, wellbeing and the minimum wage offered by the government.

In the investigation of patterns of adult mortality and its main contextual determinants, different machine learning algorithms have been employed to understand the association between the input variables and the variable of interest (probability of adult mortality, which is the probability of reaching 15 years of age and dying before completing 60 years of age). The use of machine learning methods in the social sciences enables performing science *a posteriori* by seeking patterns, finding associations, visualizing the complexity of phenomena and predicting health outcomes without considering hypotheses formulated *a priori* (without necessarily beginning with a prior theoretical foundation). This means transitioning from a hypothesis-driven approach to a data-driven approach.

The aim of the present study was to investigate associations between the probability of adult deaths in microregions of Brazil in 2010 and socioeconomic, structural, contextual and health-related factors to identify groups in situations of risk and vulnerability. This paper contributes to the literature by using a methodological approach directed at understanding patterns in data and focusing on small areas within a highly unequal and diverse country.

Understanding the heterogeneity in the distribution of the probability of adult deaths among different regions of Brazil is necessary to the development of more effective public policies, as regional differences in adult mortality reflect differences in risk factors, such as population structure, access to healthcare services, the quality of hospital care and variations in risk behaviors.

## Inequalities in Brazil

As adult mortality rates are not homogeneous in Brazil, the experience of the country does not reflect the experience of specific population groups. Mortality rates differ depending on different dimensions, such as age, race, socioeconomic status and geographic context. Geographic inequalities in mortality seem to be larger in Brazil than other countries of Latin America and the different regions of the country also have very different experiences in terms of environmental exposure, disease control, medical treatment, risk and welfare (Fenelon 2013; Brant et al. 2017; France et al. 2017).

The macroeconomic aspect is a contextual factor that regards the economic indicators of a geographic area as a whole rather than individual characteristics. Macroeconomic factors can have either a direct or indirect effect on immediate risk factors that are directly associated with health. One explanation given for the association between greater income and better health is the tendency to enhance social cohesion and reduce social divisions, whereas, contrarily, poverty is associated with social exclusion (Spijker 2004).

The most widely used measure of inequality is the Gini coefficient. Based on this index, the degree of income inequality in Brazil diminished at an accelerated pace between 2001 to 2005, going from 0.593 to 0.566. Despite this 4.6% reduction, Brazil continues to be among the most unequal countries in the world (Barros et al. 2007).

Brazilian economic development was not capable of attenuating the high concentration of wealth and the country continues to experience acute socioeconomic inequality to the present day. In 1980, 47.9% of the wealth was concentrated in the hands of the richest 10% of the population, whereas only 1% of the income was in the hands of the poorest 10% (Wood and Carvalho 1994). This concentration was maintained in 2005 despite the reduction in income inequality, as the richest 10% concentrated more than 40% of the wealth and the poorest 40% had less than 10% of the wealth (Barros et al. 2007).

Studying socioeconomic differences in a Brazilian city, Belon, Barros and Marín-León (2012) found that the highest mortality rates among adults were concentrated in areas with precarious living conditions and the mortality gradient increases with the reduction in the socioeconomic level of the city. Moreover, social inequalities in mortality were higher among the younger population compared to the older population.

## Data and Methods

### Data

#### Sources of data

The data came from the 2010 demographic census performed by the Brazilian Institute of Geography and Statistics as well as the Mortality Information System of the National Registry of Health Establishments and the Primary Care Information System organized by the Health Ministry through the Informatics Department of the Brazilian public healthcare system also for the year 2010.

The units of analysis were the 558 microregions defined by the Institute of Geography and Statistics. The microregions are groups of municipalities based on economic and social similarities. The demographic census unites information on the Brazilian population, such as schooling, income, employment, habitation and social vulnerability, in 10-year periods and has national coverage.

The Registry of Health Establishments has information on the infrastructure of healthcare services, type of care offered, specialized services, number of available hospital beds and existing capacity, with the aim of assisting in health planning. It is the official registry of the Health Ministry regarding the installed capacity and labor force of the healthcare system in Brazil considering public and private establishments. The main purposes are to register information on healthcare establishments in terms of physical and human resources and maintain the records updated.

The Primary Care Information System offers information on records of families, housing and sanitation conditions, the health situation, and the production and composition of health teams. It is the main instrument for monitoring the actions of the Family Health Program. Its purpose is to monitor and evaluate primary care and consolidate the evaluation in the three management tiers of the public healthcare system as well as monitor the actions and results of activities conducted by the teams of the Family Health Program.

#### Independent Variables

Table 1 displays the variables used in the models, the sources from which the data were obtained and a description of how the variables were used.

**Table 1:** Variables used in the model and sources Variables.

| **Variables** | **Name** | **Source** |
| --- | --- | --- |
| gdp_per_capita | Gross Domestic Products per capita | SIAB |
| income | Household Income | Census |
| higherdegree | Proportion of population with higher education (college) | Census |
| cov_garbage | Proportion of households waste collection | Census |
| urbanization | Degree of Urbanization | CNES |
| hospitalBeds | Hospital beds per 1000 people | CNES |
| cov_water | Proportion of households with piped water | Census |
| cov_sewerage | Proportion of households with sewage collection system | Census |
| white | Proportion of white people | Census |
| aging_index | Aging Index | Census |
| circsystem | Mortality Rate by Circulatory System Diseases | Census |
| child.labor | Child Labor Rate | Census |
| sex_ratio | Sex Ratio | Census |
| cov_vaccination | Vaccination Coverage | SIAB |
| cov_FHS | Coverage of Family Health Strategy | Census |
| illiteracy | Illiteracy Rate | Census |
| poverty_rate | Poverty Rate | Census |
| externalcauses | External Causes Mortality Rate | Census |
| unemployment | Unemployment Rate | Census |
| blacks | Proportion of black/brown | SIM |
| young_prop | Proportion of young people (15 to 29 years) | SIM |

**Source:** IBGE (2010); Datasus (2010).

The variables have different angles of socioeconomic, demographic and health-related characteristics in the microregions of Brazil. The column ‘Variables’ lists the names of the variables as they were used in the databank. ‘Name’ is the complete name of the variable and the source from where it was obtained.

#### Variable of interest: Probability of adult death

In Brazil, mortality studies are limited due to the lack of quality of the data and it is necessary to perform prior correction of the under-reporting of deaths so that the data reflect the actual number of deaths.

In this paper, the mortality estimate created by Schmmertman and Gonzaga (2018) was used, applying the TOPALS technique with the empirical Bayesian method to small areas for the correction of deaths. The mortality estimate in small areas has a universal problem of small populations and high sampling variability in the deaths registered as well as low mortality rates and short exposure periods. Event/exposure rates are very unstable, making the estimate of mortality patterns difficult. In such situations, death correction models must have ways to fill the gaps.

Schmmertman and Gonzaga (2018) created the entire model in the *Stan* language (Carpenter et al. 2017). For both sexes, we removed the estimated information on the distributions of mortality variables (α) and complete tables of the mortality rate logarithm of all 558 microregions. Mortality patterns are less clear in small areas. In Brazil, differences are found in mortality by specific age per cause of death and age patterns of mortality for all causes (Prata 1992; Araújo 2012).

Using the logarithm, the mortality rate by simple age (0 to 99 years) was obtained per microregion and used to calculate the probability of survival from age n to age x, p_x_(_n_p_x_ = exp(-_n_m_x_)) and the number of survivors at age x in the cohort (l_x+n_ = l_x_*_n_p_x_), thereby calculating the probability of adult death:_45_q_15_ (_45_q_15_ = l_59_ / l_15_).

After this calculation, the probability of adult death was transformed into a variable with two classes using the median probability: microregions with a probability of adult death less than 0.1406 (midpoint) were classified as P0 and the rest were classified as P50. This approach was chosen to improve the performance of the machine learning algorithms. Lustgarten et al. (2008) showed that the transformation of the continuous variable into two classes can significantly improve the classification performance of these algorithms.

### Methods

#### Machine learning

There are two major categories of machine learning methods: “supervised” and “unsupervised”. “Unsupervised” learning regards methods for finding patterns in data and reducing data (Kuhn and Johnson 2013) and is widely used to group non-labeled data based on the similarity of the characteristics, whereas “supervised” learning is adequate for predictive modeling based on the construction of relations between inputs (such as, socioeconomic characteristics) and the outcome of interest (probability of adult death).

An individual characteristic of supervised algorithms is the construction model is oriented by data, such that the data can be adjusted to complex relations automatically, which eliminates the effort in selecting variables and creating models. More specifically, machine learning algorithms can automatically detect non-linearities and non-additives.

To understand the interaction between the explanatory variables and the outcome of interest, three different types of supervised algorithms were used in the present study: *Random Forest* (RF)*, Extreme Gradient Boosted Trees* (XGBoost) and *Support Vector Machine* (SVM).

#### Pre-processing

Some transformations of the explanatory variables were necessary, as the selected variables have very different scales. For instance, vaccine coverage is a proportion (%) and GDP per capita ranges from 2,000 to 20,000.

Two transformations were performed: centralization and transformation on the same scale. To centralize an explanatory variable, the mean of the variable is subtracted from all values. As a result of centralization, the variable as a zero mean. For data to be on the same scale, each value of the variable is divided by its standard deviation. Placing data on the same scale forces the values to have a common standard deviation of 1. These manipulations are generally used to improve the numerical stability of machine learning algorithms. Some models benefit from the variables being on the same scale (Kuhn and Johnson 2013).

#### Evaluation metrics

The metrics used in the evaluation of the algorithms are generated from a confusion matrix, which is a matrix that combines both correct prognoses performed by the algorithms and incorrect prognoses. The values of the confusion matrix generated by each algorithm are used to calculate the following metrics: 1) accuracy – number of correct predictions among all predictions made by the model; 2) sensitivity – ratio of true positives to true positive plus false negatives; and 3) specificity – ratio of true negative to true negatives plus false positives.

The confusion matrix is generated by the combination of test subsets. In the present study, cross-validation in k-subsets was selected to evaluate the models. This is a procedure in which the sample is divided into k subsamples of equal size. The models are trained with k – 1 subsets (training set) and then tested with the remaining set (test set) to evaluate the classification performance of the model, iterating by means of each of the k samples. The choice of k is generally 5 or 10. k = 10 was chosen for the four algorithms analyzed in the present study. In other words, a subset of samples is used to adjust a model and the remaining samples are used to estimate the adequacy of the model. This process is repeated several times and the results are aggregated and summarized (Kuhn and Johnson 2013; James et al. 2017).

Besides the performance metrics, the importance of variables with the greatest relevance to the classification of a microregion as having a high or low probability of adult deaths was calculated to enable a better interpretation of the results. To gain a better understanding of the microregions classified incorrectly, the first eight variables with greater predictive power in the model with the best performance were chosen to compare their pattern in relation to the P0 (low probability of adult death) and P50 (high probability of adult death), contrasting the patterns found in the samples of microregions classified correctly by the algorithms and those classified incorrectly.

#### Support Vector Machine

Support Vector Machine (SVM) is one of the most common methods applied to supervised classification problems, mainly due to its excellent precision and generalization properties (e.g., Podda et al. 2018; Hsieh et al. 2016). The basic concept behind the SVM is the discovery of a hyperplane that can separate data into the number of classes, projecting data into a dimensional space by the application of a “kernel” function.

#### Random Forest

Several decision trees are constructed from data and the decisions of each tree are used to obtain a single tree (Pan et al. 2017; Nguyen 2016). With the *Random Forest* (RF) method, decision trees constructed by taking randomly resampled partitions of data and using a random subset of potential variables to inform each division of the data. By creating random samples with observations and determining possible division variables, this method introduces an element of randomization that impedes the overfitting of the model. After training, all trees are combined based on the predictions of each individual tree (James et al., 2017).

Tree-based algorithms facilitate the understanding of the results and the interactions among the variables used in the classification. The advantage of RF is the fact that this approach makes use of several trees with a random subset of resources for training and testing, leading to greater diversity and a more robust prediction.

#### Extreme Gradient Boosted Trees (XGBoost)

Gradient boosting algorithms are also based on a set of trees, but the XGBoost method begins with weak learning trees and constructs more reliable trees based on the residuals of the prediction of each previous tree. This differs from the RF approach, in which trees are independent of each other and the method depends on the random selection of variables and observation to produce impartial estimates (Nguyen 2016).

The TrainControl and Train functions of the *caret* package of the RStudio version 3.5 were used for the implementation of the algorithms. This package enables implementing numerous machine learning algorithms and the main resampling methods, including cross-validation by k-subsets, as described above.

## Results

The majority of Brazilian microregions have water and sewage coverage for more than 90% of homes. Some microregions have lower coverage rates (Table 2).

**Table 2:** Summary of Predictor Variable Statistics.

| **Variables** | **Min** | **Median** | **Mean** | **Max** |
| --- | --- | --- | --- | --- |
| gdp_per_capita | 2,867 | 12,105 | 14,105 | 77,872 |
| income | 162 | 542 | 542 | 1,665 |
| higherdegree | 0.02 | 0.07 | 0.07 | 0.24 |
| cov_garbage | 0.18 | 0.79 | 0.76 | 1.00 |
| urbanization | 22.34 | 72.72 | 72.01 | 100 |
| hospitalBeds | 0.39 | 2.12 | 2.29 | 9.34 |
| cov_water | 0.45 | 0.99 | 0.96 | 1.00 |
| cov_sewerage | 0.31 | 0.91 | 0.87 | 1.00 |
| white | 0.08 | 0.40 | 0.45 | 0.92 |
| aging_index | 8.47 | 41.82 | 42.42 | 87.76 |
| cirsystem | 8.05 | 171.45 | 169.78 | 342.55 |
| child.labor | 3.31 | 10.79 | 11.51 | 31.27 |
| sex_ratio | 88 | 99 | 100 | 114 |
| cov_vaccination | 0.56 | 0.77 | 0.77 | 1.00 |
| cov_FHS | 0.03 | 0.77 | 0.74 | 1.69 |
| illiteracy | 2.30 | 11.25 | 14.43 | 42.40 |
| poverty_rate | 0.00 | 0.04 | 0.07 | 0.30 |
| externalcauses | 0.00 | 68.99 | 70.88 | 154.87 |
| unemployment | 1.10 | 6.74 | 6.93 | 20.47 |
| blacks | 0.07 | 0.58 | 0.53 | 0.87 |
| young_prop | 0.21 | 0.27 | 0.27 | 0.32 |

Source: IBGE (2010); Datasus (2010).

An analysis of schooling indicators, such as the illiteracy rate and proportion of individuals with a university education, reveals a considerable diversity of levels; microregions have high illiteracy rates and low proportions of individuals with a higher education. The mean of individuals who studied at the undergraduate level of higher education was 8.3% in 2010, with the highest levels of education found in the southeastern region of the country (Institute of Geography and Statistics, 2010a). The mean illiteracy rate in the country was 9.6% in 2010, but with unequal distribution. The rate in the northeastern region of the country was 28% as opposed to the 5% rate in the southern and southeastern regions (Institute of Geography and Statistics, 2010a).

The socioeconomic indicators (poverty rate, child labor rate, mean household income per capita, GDP per capita and unemployment rate) also exhibited important regional differences, with a mean poverty rate of 7% for the country as a whole, reaching 30% in the microregion of Traipu in the state of Alagoas (northeastern region of Brazil). The lowest poverty rates were found in microregions of the southern and southeastern regions.

The unemployment rate had a minimum value of 1.3% of the economically active population that was without a job, with a mean of 6.9% and a maximum of 20.5%. The northern and northeastern regions had the highest concentration of microregions with higher unemployment rates. The southeastern, southern and central western regions also had microregions with high unemployment rates, but at a lower proportion compared to the northern and northeastern regions. The GDP per capita, which indicates the level of economic production of a given Brazilian microregion in 2010, had a minimum value of R$ 2,867.00 per capita, a mean of R$ 14,105.00 per capita and a maximum of R$ 77,872.00 per capita.

The GDP of the states of São Paulo, Minas Gerais and Rio de Janeiro corresponded to 50% of the GDP of the country, whereas 18 other states together accounted for only 22.2% of the GDP (Santos; Pales; Rodrigues 2014). Moreover, the lowest mean monthly income per capita (less than R$600.00) was found in the northern and northeastern regions.

With regards to health, most microregions (more than 70%) were covered by the Family Health Program and the number of hospital beds per 1000 residents was higher than two. The World Health Organization (WHO) defines the ideal number of available hospital beds as being between three and five. Thus, the mean in Brazil is lower than the WHO recommendation in the majority of microregions. The few microregions with more than four available hospital beds per 1000 residents were all located in the states of Minas Gerais, São Paulo, Rio de Janeiro, Paraná and Rio Grande do Sul.

Regarding demographic variable, the mean degree of urbanization was higher than 70%, the proportion of young individuals between 15 and 29 years of age was between 20 and 30% in all microregions and the aging index had a broad distribution. The mean aging index was approximately 45 individuals aged 60 years or older for every 100 less than 15 years of age (Closs and Schwanke, 2012).

The correlation of the predictor variables to each other was estimated. Figure 1 illustrates the correlation matrix with each predictor variable presented in pairs. Each correlation is color coded according to its magnitude. The visualization of the data in Figure 1 is symmetrical; the upper and lower diagonals display identical information. Dark blue indicates a strong positive correlation and dark red is used for strong negative correlations. Tones closer to white indicate a lack of correlation between predictors.

**Figure 1:**
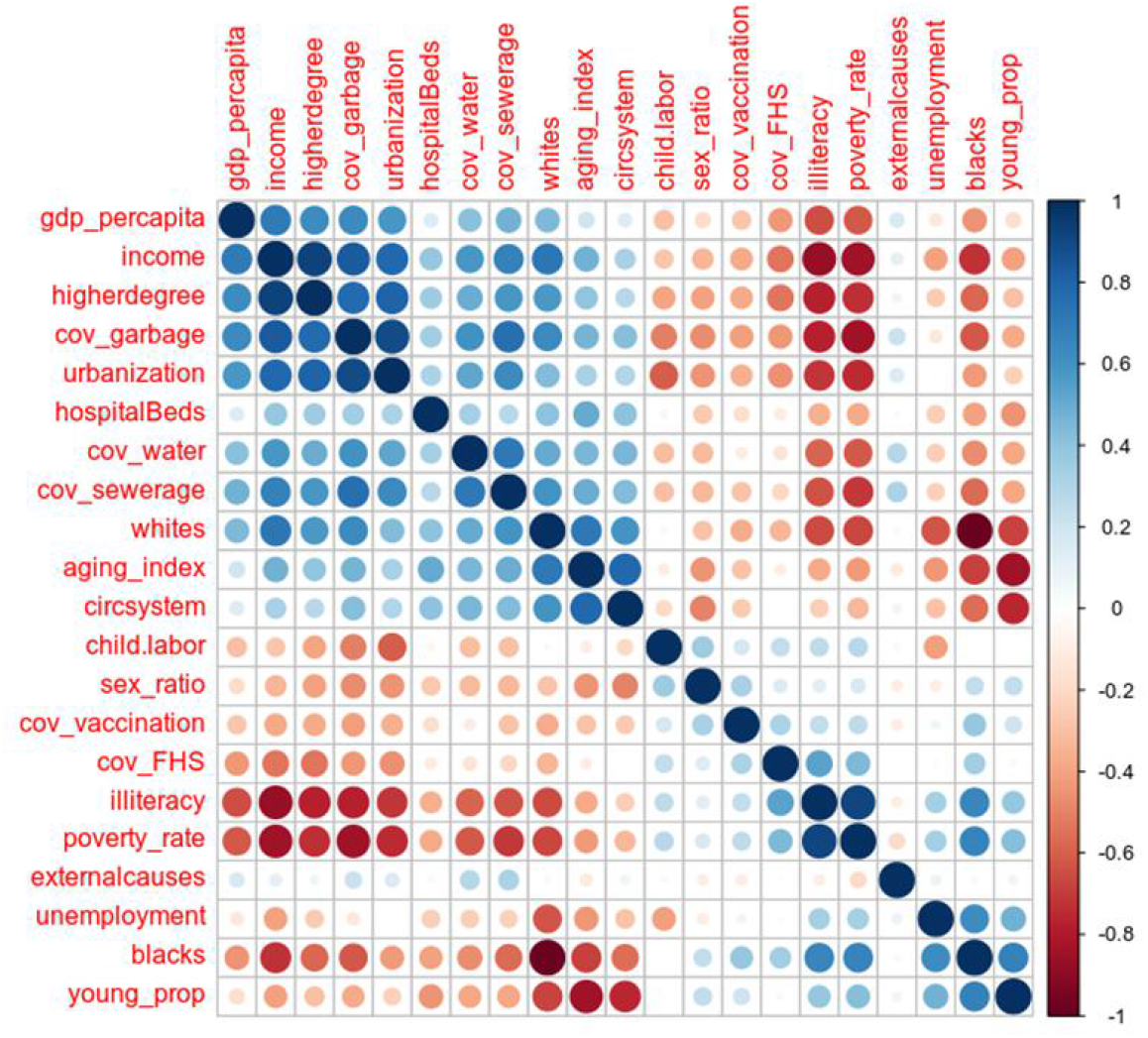
Correlation Matrix.

There are some small blocks of strong correlations (dark blue and red) near the main diagonal. One of the blocks is in the left corner, where variables linked to the socioeconomic characteristics of the microregions are found. The proportion of individuals with a university education was positively correlated with mean household income, trash collection coverage and the degree of urbanization. Moreover, poverty and illiteracy rates were positively correlated to each other. The mortality rate due to the circulatory diseases and the aging index were also correlated.

Among the negative correlations (dark red), the proportion of black/brown individuals was correlated with the proportion of white individuals and the illiteracy rate was correlated with both mean household income. The findings indicate strong correlations between black/brown individuals and income, the illiteracy rate and the poverty rate. A strong positive correlation was also found between the literacy and poverty rates.

Likewise, strong negative correlations were found between mean income per capita and both the illiteracy and poverty rates, whereas strong positive correlations were found between a university education, trash collection coverage and the degree of urbanization. The proportion of whites was negatively correlated with the proportion of youths in the population and a positive correlation was found between the proportion of youths and the proportion of blacks (black and brown individuals). These correlations between youths, aging and the proportion by skin color/race may be associated with the fact that the proportion of whites over 65 years of age is 57% in Brazil, whereas the proportion of blacks in the same age group is 42% (Brazilian Institute of Geography and Statistics 2010a; Oliveira; Thomaz; Silva 2014).

The performance of the models was measured by the aggregate of 10 resamplings using the cross-validation method, dividing the data into 10 subsets.

displays the confusion matrix for all three classifiers analyzed. Accuracy was 77.6% for XGBoost, 76.2% for SVM and 74.9% for RF. Sensitivity was 79.2% for XGBoost, 77.1% for SVM and 72.8% for RF. Regarding specificity, RF had the best result (77.1%), followed by XGBoost (75.9%) and SVM (75.3%). Therefore, XGBoost had the best overall performance, followed by SVM and RF. For the subsequent analyses, accuracy is the most important measure, followed by specificity. Accuracy regards the number of times a class is predicted correctly in relation to all classes discriminated. Specificity is an important measure, as the results should contain fewer false positives to provide a more effective classification and interpretation. Accuracy is a measure that indicates how well a model discriminates unseen data, meaning that it is able to reveal the performance of a model in classifying a given microregion as having a low or high probability of adult deaths.

**Figure 2:**
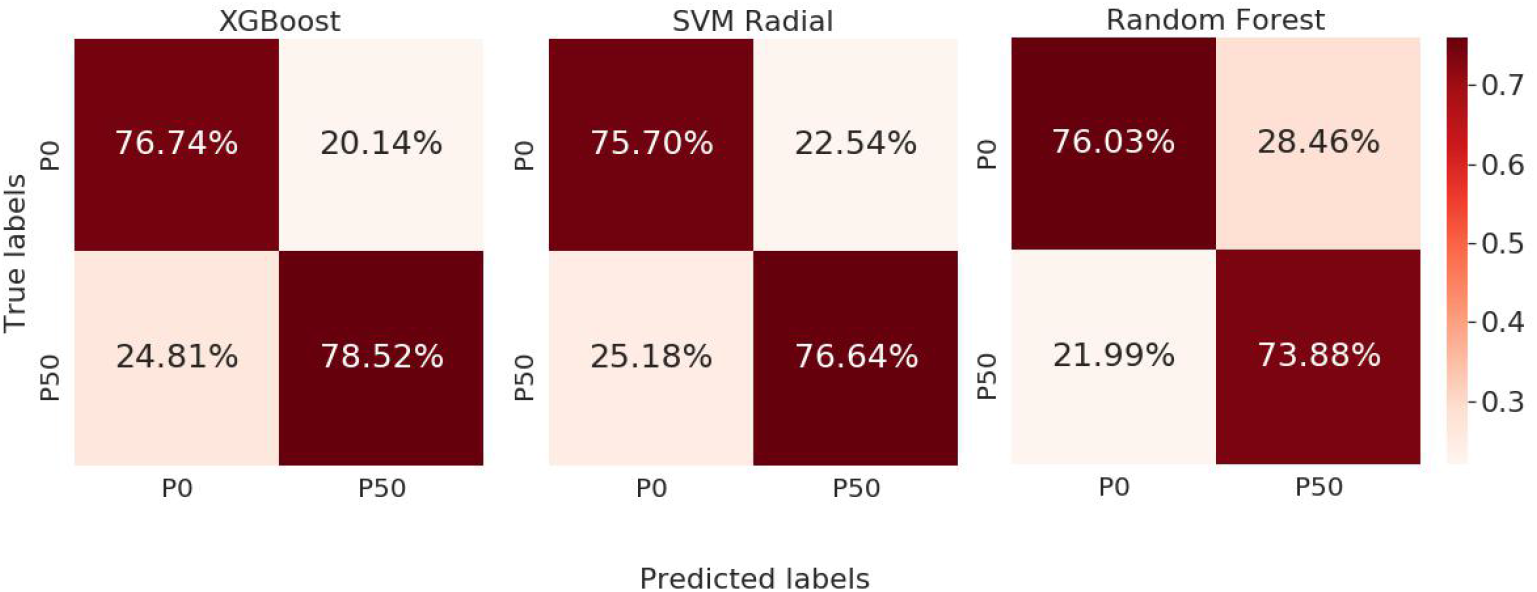
Confusion matrix for evaluated classifiers. **Source:** IBGE (2010); Datasus (2010).

## Importance of variables

The importance of variables is a measure that indicates those with the greatest relevance to the models. As XGBoost had the best performance among the three models, it was chosen to analyze the importance of the variables. Importance is calculated using the accuracy of each tree and recalculated by permutating each predictor variable. Variables with high predictive power have higher values and those with low predictive power have lower values.

Figure 3 illustrates the importance of each determinant generated by the model, demonstrating the predictive power of each variable regarding the P50 classification (high probability of adult deaths), which is a probability greater than 0.1406 (midpoint of the probabilities of adult deaths). The mortality rate due to external causes was the most important variable, the unemployment rate was the second most important variable and the sex ratio was ranked third. The variables with the least predictive power were the proportion of individuals with a university education, the number of available hospital beds per 1000 residents, coverage by the Family Health Program and GDP per capita.

**Figure 3:**
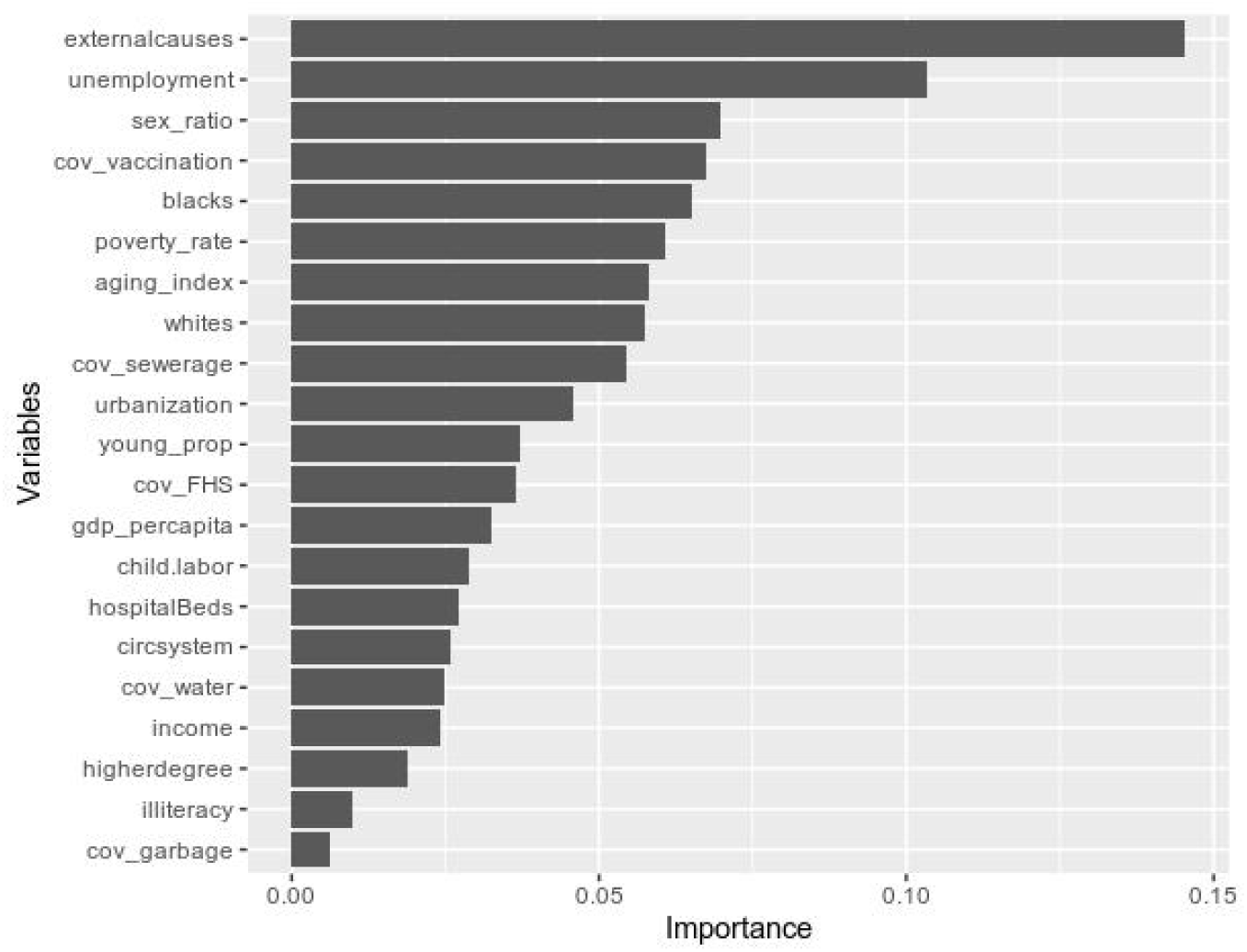
Feature Importance.

Regarding accuracy, 25% of the microregions were incorrectly classified by the algorithms. These microregions are in red in the map below (Figure 4). The question arises as to the reason for the incorrect predictions. Based on some of the characteristics of these microregions, the algorithm classified some as having a low probability of adult deaths when they actually had a high probability or vice versa.

**Figure 4:**
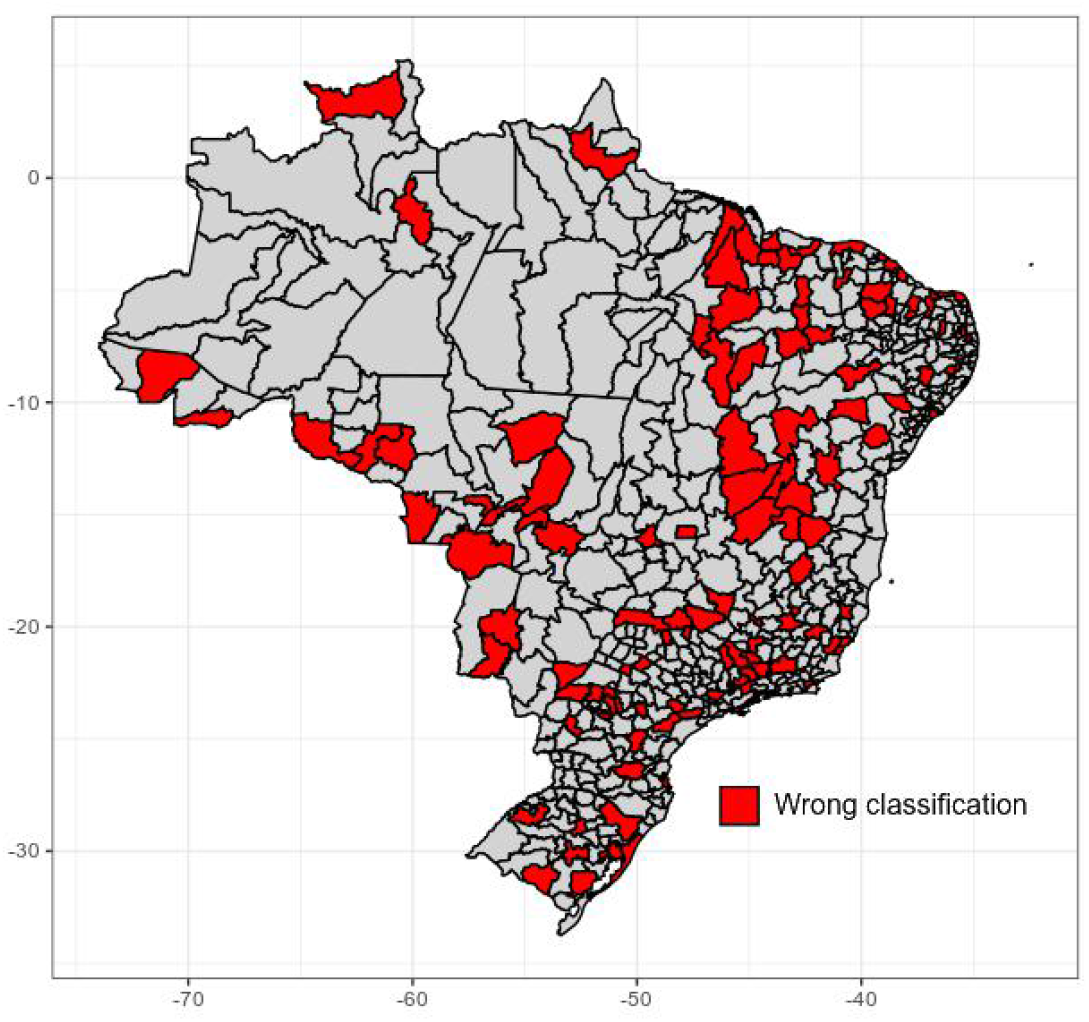
Brazil’s Map with microrregion wrongly classified by XGBoost model in red. **Source:** IBGE (2010); Datasus (2010).

Figure 5 displays the selected variables in relation to the microregions classified correctly by the models. Some patterns are found among microregions with high adult mortality. For instance, considering the pink curve, which represents the distribution of the P50 class, the proportion of blacks is more to the right, denoting higher values in microregions with high adult mortality. The same is true for unemployment. In contrast, the aging index has lower values in these microregions.

**Figure 5:**
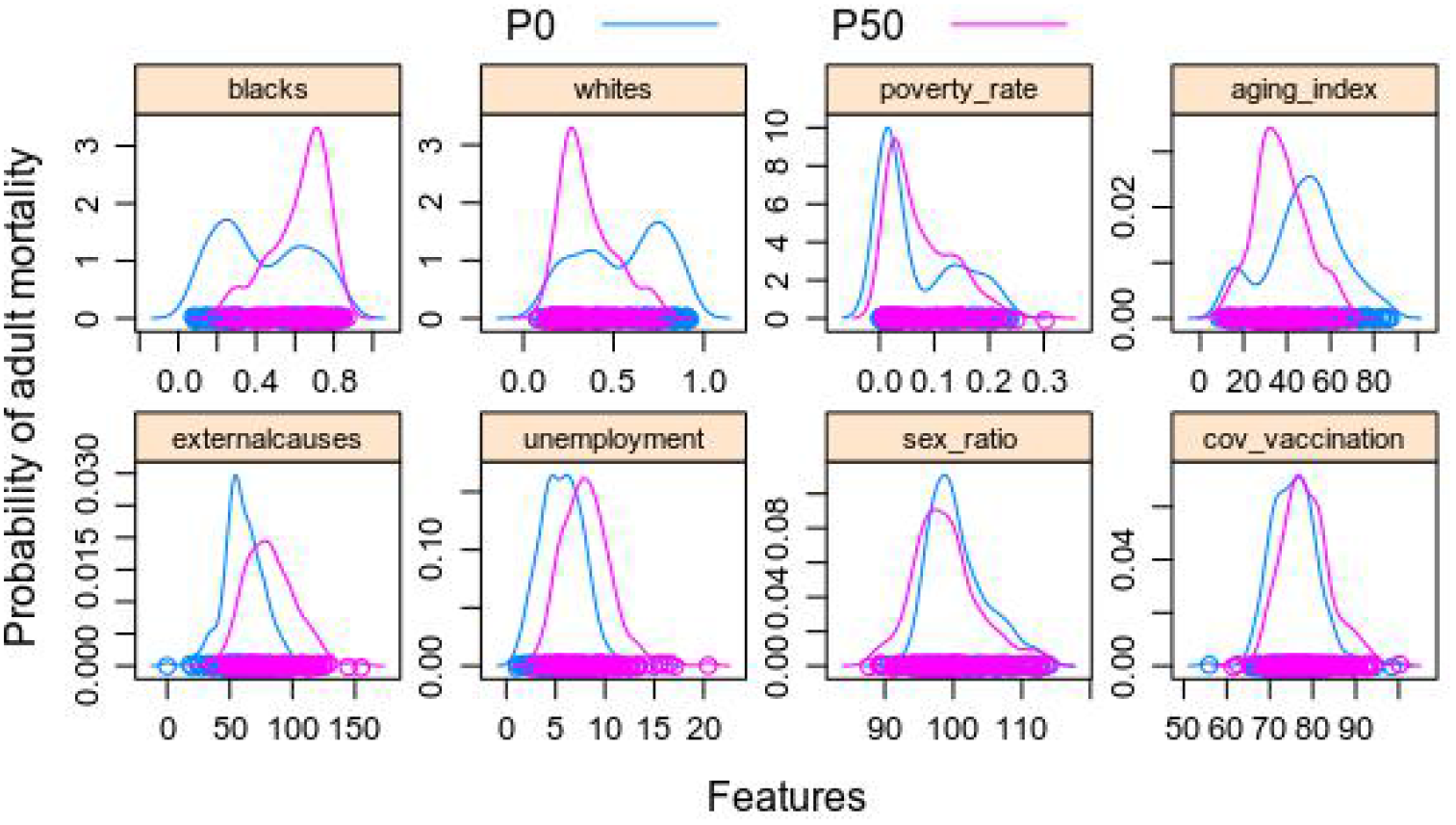
Sample with the right classifications made by the XGBoost.

The microregions classified incorrectly by the XGBoost model (Figure 6) have a different profile. The proportion of blacks (black and brown individuals) has a distribution with smaller values, the aging index is more to the right for microregions with higher adult mortality and the mortality rate for external causes is lower compared to microregions with low mortality.

In this analysis, microregions classified incorrectly with regards to adult mortality (Figure 6) have a different profile from the majority of microregions classified correctly (Figure 5). One may suppose that other variables not considered in the models make these microregions have different adult mortality compared to the more common profile found in the model for classifying microregions as having a high or low probability of adult deaths.

**Figure 6:**
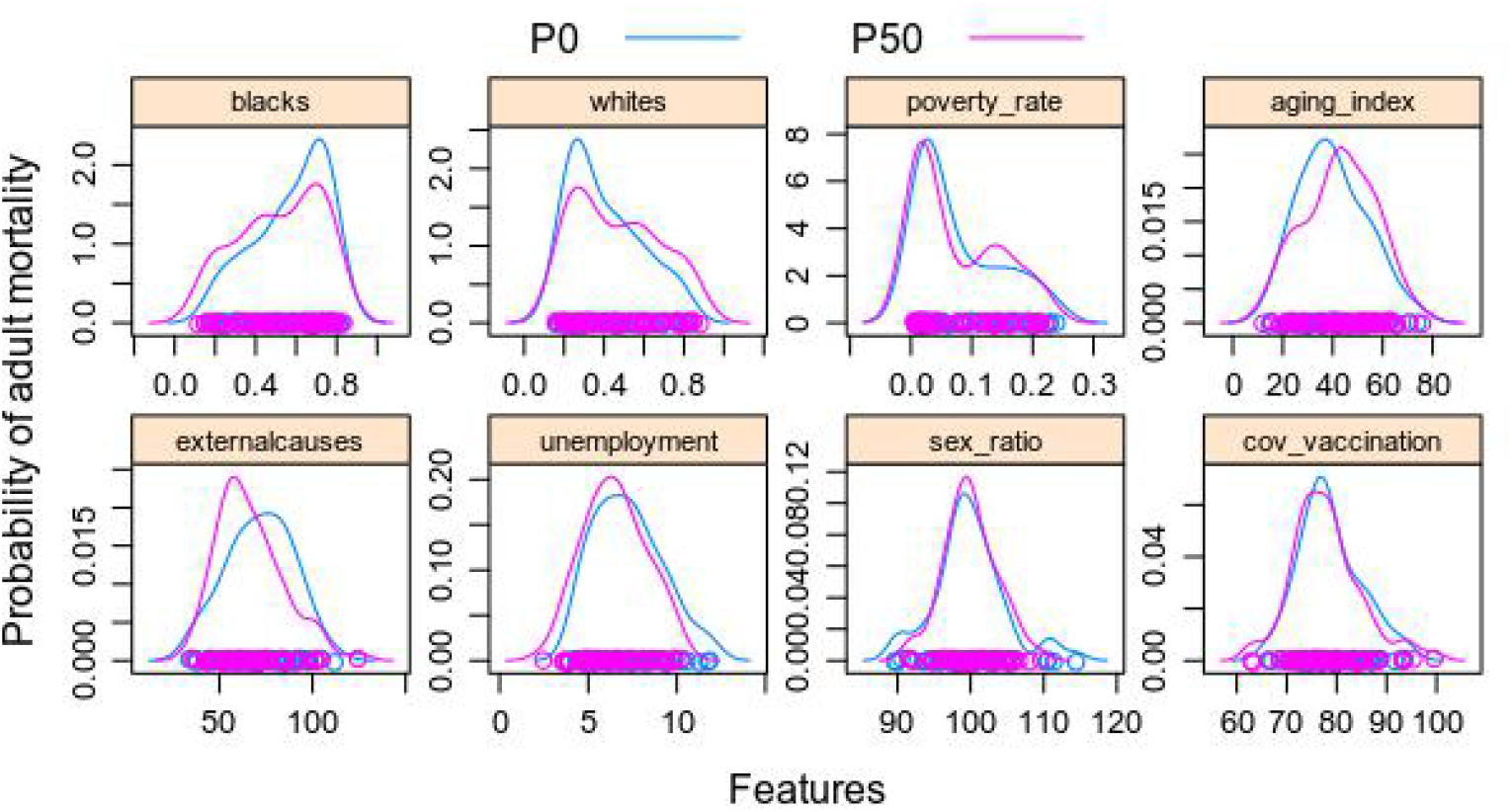
Sample with the wrong classifications made by the XGBoost. **Source**: IBGE (2010); Datasus (2010).

## Discussion

In the present investigation, the mortality rate due to external causes, the proportion of black/brown individuals and the proportion of whites were among the main variables with predictive power regarding adult mortality. These findings are in agreement with data described by Araújo et al. (2009), who concluded that skin color exerts an impact on the potential lost years of life due to external causes, as the number of years lost for this reason was 8.8-fold higher in the population of black and brown individuals in Brazil.

In a study conducted with municipalities in the state of São Paulo, mortality between 15 and 44 years of age was twofold higher in areas with poorer social and environmental conditions compared to areas with better conditions (Stephens et al. 1994). In 2008, Campos et al. (2015) investigated the profile of deaths by age group, sex and region and found that external causes accounted for 10% of the total burden. Homicides and violence were the main reasons for the lost years of life, accounting for 38.8%. In the northern and northeastern regions of the country, these figures were 43.9% and 46.6%, respectively, followed by traffic accidents (36% of lost years of life).

Homicide rates increased in the northern and northeastern regions between the year 2000 and 2010, whereas the rate diminished in the southeastern region in the same period (Campos et al. 2015). The increases in these regions may be associated with the accelerated urbanization process, leading to new economic centers without the adequate structuring of public safety and infrastructure policies (Waiselfisz, 2014; Ipea and FBSP, 2018). Another reason may be related to the age structure. A study conducted in the state of São Paulo reports that nearly half of the reductions in homicide rates in recent years could be explained by the increase in the older population and decline in the young population, which is more vulnerable to the impacts of violence (Mello and Schneider 2007; Brasil 2015b).

In a study on socioeconomic inequalities in the United States, Bosworth (2018) points out that racial differences are strong factors of the disparity found in mortality rates at most ages, but the interaction between the role of race and socioeconomic status remains the object of debate. Regarding inequalities in health, Williams (2016) reports the occurrence of considerable racial differences in the United States regarding the quality of medical treatment even after adjusting for access to services, disease severity and socioeconomic status.

Studies conducted in the United States and Brazil show that areas where the population is made up of a large proportion of blacks have unfavorable socioeconomic and infrastructure indicators. This segregation contributes to considerable differences in terms of schooling, employment, income and health, limiting possible opportunities for social ascension and increasing the number of homicides and violence in this population group (Brasil 2005; Ipea 2011; Nuru-Jeter and Laveist 2011; Fiorio et al. 2011; Petrucelli and Saboia 2013; Oliveira-Thomaz and Silva 2014; Cunninghan et al. 2017). The classification of the algorithm shows that race tends to predict increased health risks and mortality independently of economic status, which is in agreement with data reported by Lovell (1999).

Unemployment was the variable with the second most predictive power, which validates the vast literature documenting the strong associations between disadvantage in the job market and both health and mortality (Iversen et al. 1987; Moser et al. 1987; Roelfs et al. 2011, Queiroz et al. 2017). High unemployment rates are related to low growth and economic development. According to Gerdtham and Johanneson (2003), the association between unemployment and the increase in mortality may be mediated by psychological malaise (such as depression) and unhealthy behaviors (such as substance abuse). In contrast, employment has been associated with positive characteristics, such as greater self-esteem and independence.

Queiroz et al. (2017) found that high levels of unemployment were associated with a greater probability of adult deaths in Brazil. In a longitudinal study conducted in Sweden, Montgomery et al. (2013) found that among men aged 34 to 38 years and those aged 45 to 49 years, the risk mortality related to unemployment is greater in the older group and among more qualified individuals. With the increase in age, the occurrence of unemployment may be unexpectedly harmful. A lack of adaptation to social adversity, such as losing one’s job, can lead to an increased risk of stroke. This highlights the greater vulnerability of older workers in times of recession and higher unemployment rates (Montgomery et al. 2013).

At the other end of the spectrum, unemployment among the young population in the transition to adulthood (18 to 24 years of age) is related to higher risks of general mortality, homicide and all-cause mortality after controlling for sex, race and education (Davila et al. 2010). The literature on injury rates among young workers and mortality trends among individuals aged 15 to 24 years in the USA indicate that young adult males have a greater probability of dying, which may be due to more aggressive, riskier behaviors.

Clemens, Popham and Boyle (2015) report that unemployment is more likely to occur among individuals with a lower socioeconomic status. Moreover, it may be that the harmful effects to health associated with poverty and disadvantage prior to unemployment are more responsible for increasing the risk of mortality than the effects of unemployment *per se*. After a health-based comparison, the authors found an 85 and 50% increase in the risk of mortality for men and women, respectively, who were registered as unemployed for ten years in comparison to those who remained at the job. Work that is not recovered within a short period of time results in a loss of income and can rapidly impose economic hardship. Acute stress and material deprivation tend to emerge among many displaced workers after the initial loss of employment as uncertainty increases regarding immediate and future life prospects, with greater consequences among workers who are more vulnerable due to social and health-related circumstances (Garcy and Vagero 2012).

Vaccine coverage in a microregion had high predictive power. This coverage is a public health intervention and greater coverage rates are associated with better health indicators and a lower mortality rate. Population segments with poorer socioeconomic indicators generally have lower vaccine coverage rates. This association can be understood as a living condition aspect that hinders access to vaccines. Therefore, a lower socioeconomic status can lead to a lower offer of services and greater difficulty in gaining access to a particular health intervention, such as vaccination campaigns (Travassos and Martins 2004; Moraes and Ribeiro 2008).

In the context of adult age group, a survey by the Brazilian Health Ministry through the National Immunization Program showed that coverage in the 20-to-59-year-old age group with vaccines recommended for adults (hepatitis B, yellow fever and adult triple and double viral vaccines) is far below the ideal (Brasil 2015a). The main vaccination campaigns target children, pregnant women and seniors and there is no measure of coverage for the adult population. Although the survey used immunization coverage for children, the different models were able to find an association between vaccination coverage in the microregion and adult mortality, which may be related to the vaccination insufficiency in the adolescent and adult groups shown by the survey cited (Brasil 2015a).

The analyses revealed that all models were capable of significantly discriminating the lower probabilities (P0 class) and higher probabilities (P50 class) of adult deaths using a set of socioeconomic, demographic and health-related characteristics per microregion, demonstrating the importance of these characteristics regionally as indicators and determinants of mortality in the 15-to-59-year-old age group.

The microregions classified incorrectly indicate that beyond the variables of greatest influence that helped correctly classify the majority of microregions, there may be other characteristics that influence whether a microregion has a high or low probability of adult deaths, such as particular public policies and access to healthcare services.

Mortality rate due to external causes was the variable with the highest predictive power regarding the classification of a high or low probability of adult deaths in a microregion. Homicide is the major cause of death among 15-to-29-year-olds, accounting for 60% of deaths due to external causes in this age group. In the 30-to-44-year-old and 45-to-59-year-old age group, homicide rates diminish and there is a greater proportion of traffic accidents as external causes (Ferreira and Araújo 2006).

The importance of the proportion of black/brown individuals and proportion of whites in microregions to the prediction of adult mortality is evidence that health inequalities in the Brazilian population are strongly related to the social-historical construction that led Brazilian society to a division by population subgroups based on race/skin color and points to a lack of equity. In other words, a greater proportion of black and brown individuals determines a higher probability of adult deaths. However, the following questions arise that cannot be answered by the analyses performed in this paper: Does being white in a microregion with a greater proportion of blacks increase the probability of adult death for this white individual? Likewise, does being black in a microregion with a greater proportion of whites diminish the probability of adult death for this black individual?

Regarding inequalities between the sexes in terms of adult deaths, the higher mortality rate among men is related to exposure to risk factors and situations throughout life, such as unsafe working situations, and harmful behaviors, such as greater use of alcohol, cigarettes and drugs, as well as the frequent exposure to risks of accidents and violence.

## Limitations and future analyses

The cross-sectional design (considering a single point in time) is the major limitation of the present study, as this design does not enable establishing relations of causality or drawing more in-depth conclusions regarding these relations. Future studies should disaggregate the probability of adult death separately by sex and smaller age groups than the large span of 15 to 59 years. Particular factors may exert different influences between the sexes and among the 15-to-29-year-old, 30-to-44-year-old and 45-to-49-year-old age groups separately. Moreover, analyzing the probability of adult deaths stratified by sex can help determine whether contextual factors exert the same influence on adult mortality in men and women. The methods of this study could also be extended to include grouped modeling techniques, combining the results of different models to create a model that considers the different elements of the data that each algorithm identified as important. The present investigation is a first step in this direction.

## Conclusion

The approach employed in the present study involving the use of contextual factors enables the determination of certain relations between social structure and health that is not possible on the individual level. This study sought to understand why adult mortality is higher in some microregions than others and why its distribution is uneven. We also sought to understand why some microregions were classified incorrectly by the algorithm. The resources of a microregion can determine the quality of life of individuals. Therefore, social context exerts a considerable influence on the determination of health conditions. Moreover, the analysis of the predictions made incorrectly by the algorithm enables us to understand that characteristics differ in the majority of microregions.

## Data Availability

Dataset will be made public after paper been published in a journal.

## Notes

### Competing Interest Statement

The authors have declared no competing interest.

### Funding Statement

This study was financed in part by the Coordenacao de Aperfeicoamento de Pessoal de Nivel Superior, Brasil, CAPES. Finance Code 001

### Author Declarations

This paper uses publicly available data (SIM, CNES and demographic census) and was deemed exempt from approval from a human research ethics committee.

